# The ABCD-GENE score influences vascular event rates in both users of clopidogrel and aspirin, as well as non-users of either drug in a population-based cohort study

**DOI:** 10.1101/2023.08.06.23293732

**Authors:** Kaavya Narasimhalu, Ernst Mayerhofer, Livia Parodi, Marios K Georgakis, Deidre Anne De Silva, Jonathan Rosand, Christopher D Anderson

## Abstract

**Background and Objectives:** Clopidogrel is an antiplatelet used in both primary and secondary prevention of cardiovascular diseases. It is a prodrug, requiring *CYP2C19* for its metabolism to the active metabolite. The ABCD-GENE score, combining clinical attributes (age, body mass index, chronic kidney disease, diabetes mellitus), with genetic information (presence of 1 or 2 loss of function (LOF) alleles in the *CYP2C19* gene) has been shown to identify patients with higher risk of recurrent cardiovascular events in high-risk populations undergoing percutaneous coronary intervention. We aimed to determine if the ABCD-GENE score or LOF alleles were associated with an increased risk of vascular events among clopidogrel users in a general population.

**Methods:** We conducted a population based cohort study with UK Biobank’s primary care prescription records to identify clopidogrel users. ABCD-GENE scores were calculated with closest available data from the first date of clopidogrel prescription. The number of LOF alleles present, and the clinical component ABCD, were separate exposures. The outcome of interest was a composite endpoint of vascular events comprised of myocardial infarction, ischemic stroke, and death due to either of these. We performed Cox proportional hazards models with clopidogrel as a time varying exposure to predict hazards of these outcomes. In order to determine the drug specificity of these exposures, the analyses were repeated in aspirin users, and in non-users of either aspirin or clopidogrel.

**Results:** Among 11,248 clopidogrel users, 3,365 (30%) developed a vascular event over a mean follow-up of 5.95±3.94 years. ABCD-GENE score ≥10 was associated with an increased risk of vascular events (HR 1.13, 95% CI 1.03-1.23). In aspirin users, and in non-users of either aspirin or clopidogrel, the ABCD-GENE score was also associated with increased risk of vascular events. In clopidogrel users, aspirin users, and non-users of either drug, the ABCD score was associated with increased risk of vascular events. The presence of two *CYP2C19* LOF alleles was associated with an increased risk of vascular events in aspirin and non-users but not in clopidogrel users.

**Discussion:** In this population-based cohort study, the ABCD-GENE score was associated with an increased risk of vascular events in clopidogrel users, aspirin users, and in non-users of either drug. The clinical component, ABCD was also associated with an increased risk of vascular events in all three groups. This suggests that the ABCD-GENE score is not specific to clopidogrel users in identifying persons at high risk of vascular events in a general sample with low baseline *CYP2C19* LOF allele frequency.

## Introduction

Clopidogrel is a widely used drug in secondary prevention of MI and stroke. Clopidogrel is a prodrug that in its active form inhibits P2Y_12_^1^. *CYP2C19* plays a major role in the activation of clopidogrel, and users with loss of function (LOF) mutations in *CYP2C19* have less of an antiplatelet effect due to insufficient inhibition of platelet reactivity by clopidogrel’s active metabolite^2^. In addition to genetic factors, clinical factors, including age, body mass index (BMI), chronic kidney disease, and diabetes mellitus have been shown to associate with high on-treatment platelet reactivity (HPR) levels while on clopidogrel^3,4^.

The ABCD-GENE score^5^, a clinical score that incorporates these clinical and genetic factors that affect clopidogrel metabolism, was developed in a high risk cohort of patients undergoing percutaneous coronary intervention (PCI) with stable angina or myocardial ischemia who were treated with drug eluting stents that required dual antiplatelet therapy for 12 months. It has been shown to identify people at an increased risk of adverse ischemic events following PCI by predicting HPR while on clopidogrel ^6,7^. This score has been validated in high-risk coronary intervention groups and in ischemic stroke patients^8^. However, the applicability of this score in the general population who may be prescribed clopidogrel for lower-risk indications has not been established.

In this population-based study, we aimed to determine whether the ABCD-GENE score was able to identify clopidogrel users at an increased risk of vascular events, and whether this predictive capacity was specific to clopidogrel users compared with aspirin users or individuals on neither drug. We also aimed to determine whether the clinical component alone, or number of *CYP2C19* LOF mutations were associated with any vascular event.

## Methods

### Participants

We performed our investigation using the UK Biobank (UKB)^9^, a population-based cohort of over half a million participants who were aged 37-73 at baseline and recruited from 2006-2010 in 22 assessment centers across the UK. The UKB administered a wide range of assessments at baseline and genotyped most of their participants. Once a participant consented for this study, their prospective and retrospective medical records in the National Health System were included in the study data. A subset (approximately 45%) of the UKB participants had primary care data included in their data collection. The UKB has institutional review board approval from the Northwest Multi-Center Research Ethics Committee (Manchester, UK). All participants provided written informed consent. We accessed the data following approval of an application by the UKB Ethics and Governance Council (Application No. 36993).

### Clopidogrel Users

We identified users of clopidogrel from the primary care prescription data, which captures drug prescriptions between 1978 to 2018. The details of the pipeline for extracting medication data have been previously published^10^. Briefly, all available ever-approved formulations of clopidogrel were gathered by using international nonproprietary names (INN), former and current trade names in the UK (via the National Health Service Dictionary of Medicines and Devices [DM+D] browser, https://services.nhsbsa.nhs.uk/dmd-browser/search), and their associated DM+D and British National Formulary (BNF) codes (**Supplementary Table S1**). Then, all primary care prescription data were searched for these formulations, and the first ever date of clopidogrel prescription was noted. Clopidogrel use was treated as a time varying exposure, with participants considered exposed for the duration of the prescription from the primary care data alone. We did not control for concomitant use of other antiplatelet or anticoagulation.

### Derivation of the Exposures

We determined the ABCD-GENE score of each participant at the time of first clopidogrel prescription. *CYP2C19* alleles of *2 (rs4244285), *2B (rs17878459), and *3 (rs4986893) were considered LOF alleles^11^. Age was determined based on the date of the first clopidogrel prescription. BMI was derived from either the baseline UKB interview, or from the primary care data, whichever was closer to the prescription of clopidogrel (mean 1.25± 4.10 years from clopidogrel prescription). Chronic Kidney Disease (CKD) was defined as a glomerular filtration rate of less than 60ml/min based on the CKD-EPI definition^12^, and was derived from either the baseline UKB interview, or from the primary care data, whichever was closer to the prescription of clopidogrel (mean 50±375 days from clopidogrel prescription). Diabetes Mellitus status was determined based on the baseline interview’s age of diabetes diagnosis, or if unknown at baseline, determined by primary care data as close to the date of first clopidogrel prescription as possible (1.16±5.4 years from clopidogrel prescription). Search codes used to define these clinical variables from the primary care data are summarized in **Supplementary Table S2**.

Based on the published ABCD-GENE scoring system^5^, older age and higher BMI were weighted with scores of +4 each, while diabetes and CKD were weighted with scores of +3 each. Participants with one LOF allele scored +6 while those with two LOF alleles scored +24 **(Supplementary Table S3)**. The resulting ABCD-GENE score was dichotomized at less than 10, or greater than or equal to 10 based on the original publication. We also assessed the clinical component of the score (ABCD) as an ordinal variable and the number of LOF alleles as a categorial variable.

### Outcome Ascertainment

UKB participants’ records have been linked with inpatient hospital codes, primary care data, and a death registry for longitudinal follow-up. Outcome events were derived using International Classification of Diseases (ICD) 10 codes (**Supplementary Table S4**) for ischemic stroke (IS) and myocardial infarction (MI) that were aligned with the diagnostic algorithm in the UKB (https://biobank.ndph.ox.ac.uk/showcase/ukb/docs/alg_outcome_main.pdf). The primary outcome was a composite endpoint of first IS, MI, or death from IS or MI after prescription of clopidogrel. In order to minimize capture of clinical events for which clinicians made the decision to start clopidogrel, IS or MI diagnoses within two weeks before or after the prescription of clopidogrel were not counted as meeting the primary or secondary outcomes.

### Sensitivity analyses

In order to determine whether the effect of the ABCD-GENE score was specific to clopidogrel users, additional analyses were undertaken in aspirin users with primary care data in the UKB who did not concurrently receive clopidogrel, as well as non-users of either clopidogrel or aspirin in the UKB. For aspirin users, as with clopidogrel, the ABCD-GENE score was calculated as close to possible to the date of first aspirin prescription. For non-users of either clopidogrel or aspirin, the baseline UKB data was used to derive the ABCD-GENE score. Aspirin use was calculated using primary care prescription data in an analogous fashion to the procedures reported above for calculating clopidogrel use.

### Data availability

UKB participant data are available through the UK Biobank after approval of a research proposal.

### Statistical Analyses

Demographic characteristics of the study population were compared between clopidogrel users of ABCD-GENE score of ≥10 and <10 by chi-squared, t-test, as appropriate. Multivariable Cox proportional hazards models controlling for age, sex, and ethnicity were performed to determine whether there was an association between the ABCD-GENE score, ABCD, and the LOF status and vascular events. Participants entered the Cox models at the start of the first prescription of clopidogrel and were censored at the time of an outcome, death, or at last known UKB update. Clopidogrel use was controlled for as a time varying exposure in all multivariable models. Analyses were also repeated in aspirin users and non-users of either aspirin or clopidogrel separately. Aspirin users entered the Cox models at time of first aspirin prescription, and non-users of either drug entered at the baseline UKB visit. Aspirin use was controlled for as a time varying exposure in all multivariable models in aspirin users. Analyses were performed on Stata version 16.1 and significance was determined as a two-tailed alpha of 0.05.

## Results

Of the 502,411 UKB participants, 15,209 had no genetic data available, and 275,104 had no primary care data available. Participants with primary care data available were slightly older, more likely to be female, and more likely of European ethnicity **(Supplementary table S5)**. Within the remaining 212,098 participants, 11,248 had been prescribed clopidogrel **(Figure 1)**. Of these, 3,575 experienced hospitalizations for a vascular event after the prescription of clopidogrel, of which 2,968 experienced a MI and 607 an IS.

**Figure 1:**
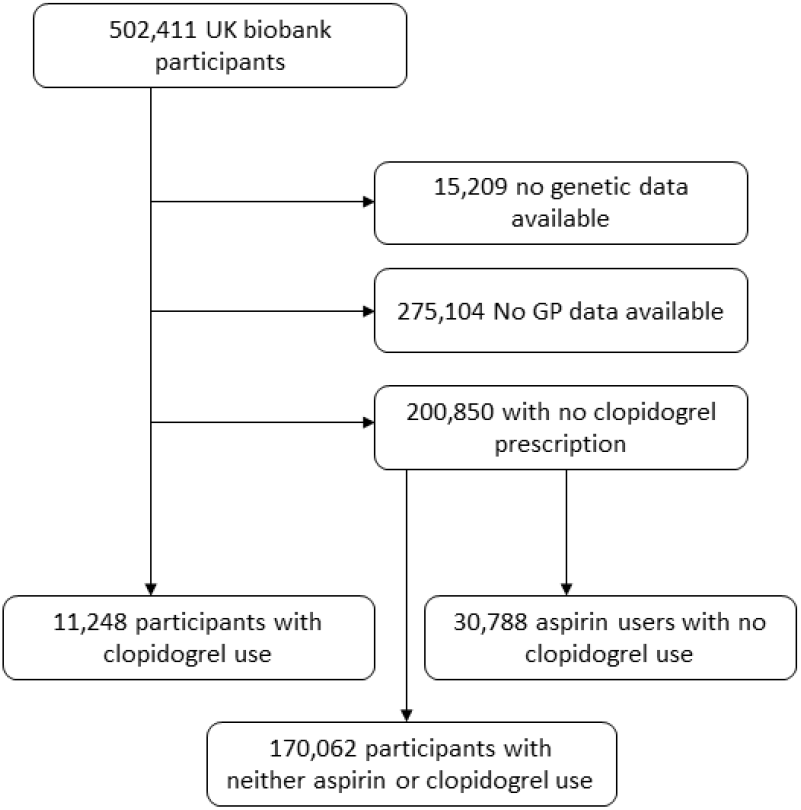
Derivation of study participants

Of the remaining 200,850 participants, 30,788 were aspirin users and the remaining 170,062 were nonusers of aspirin and clopidogrel. Demographic characteristics of clopidogrel users, aspirin users, and non-users of either drug are summarized in **Supplementary Table S6**. Briefly, non-users of either drug were significantly younger and more likely to be female than clopidogrel or aspirin users. There were no significant differences in the distribution of LOF alleles amongst clopidogrel users, aspirin users, or non-users of either drug.

The raw distribution of the ABCD-GENE score amongst users of clopidogrel is summarized in **Figure 2**. The demographic characteristics of clopidogrel users stratified by the ABCD-GENE score ≥10 or <10 are summarized in **Table 1**. Participants with an ABCD-GENE score ≥10 were less likely to be of European ancestry, and more likely to have at least one LOF allele.

**Table 1:**
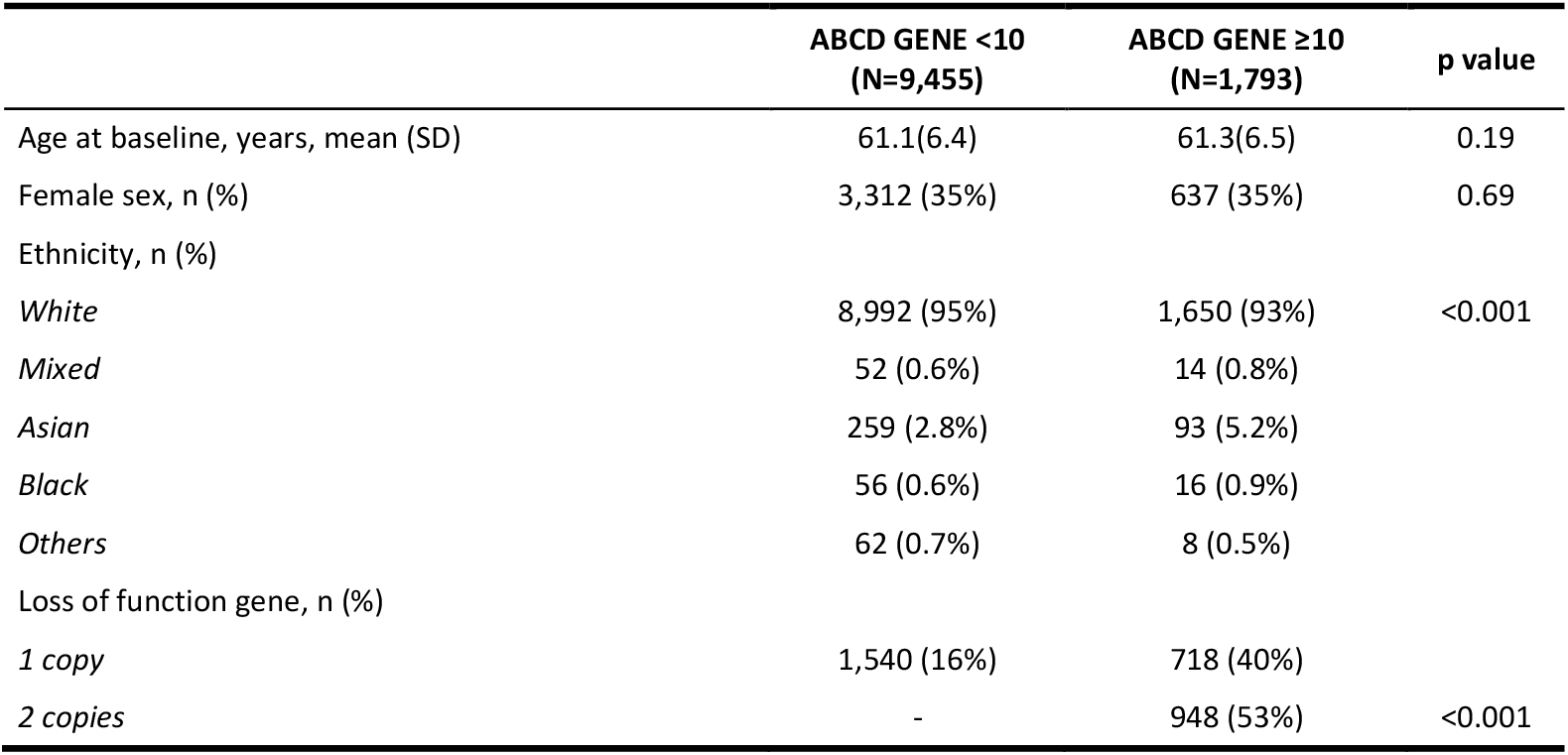
Demographic characteristics of clopidogrel users, stratified by ABCD GENE scores.

**Figure 2:**
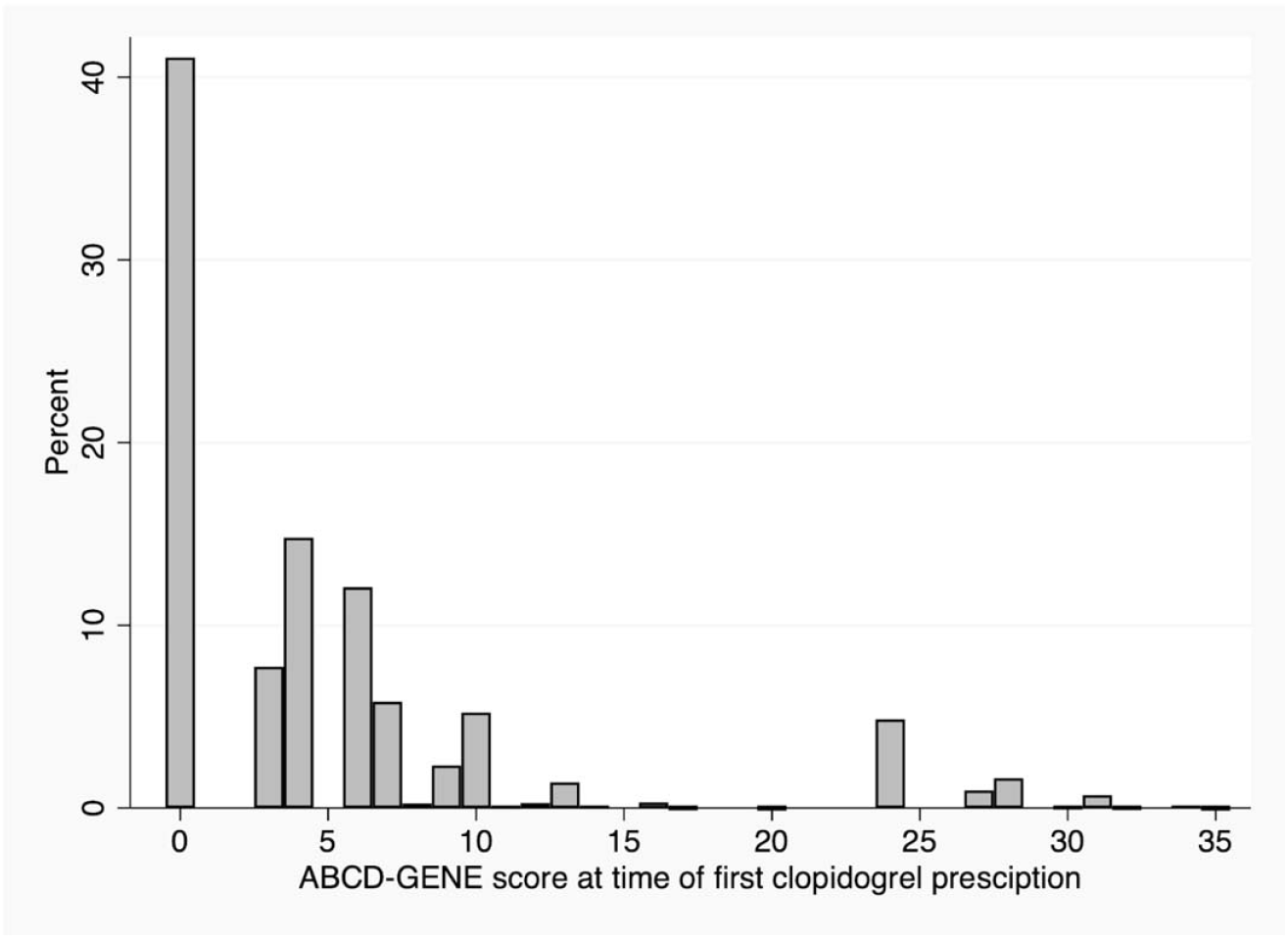
Distribution of the raw scores of the ABCD GENE score amongst clopidogrel users.

In multivariable analyses, an ABCD-GENE score of ≥10 was associated with increased risk of vascular events (**Table 2**) in users of clopidogrel, aspirin, and non-users of either medication. The ABCD score was also associated with an increased risk of vascular events in clopidogrel, aspirin, and non-users of either medication. Participants with two LOF alleles were at increased risk of vascular events amongst aspirin users and non-users of either medication, but not in clopidogrel users (**Table 2**).

**Table 2:** Multivariate Cox regression models predicting vascular events in clopidogrel users, aspirin users, and non-users of either drug.

|  | Clopidogrel users<br>N=11,248 |  | Aspirin users<br>N=30,788 |  | Non-users of either drug<br>N=170,062 |  |
| --- | --- | --- | --- | --- | --- | --- |
|  | HR | 95% CI | HR | 95% CI | HR | 95% CI |
| ABCD GENE Score $\geq 10$ | **1.12** | **1.02 – 1.23** | **1.15** | **1.06 – 1.25** | **1.20** | **1.09 – 1.31** |
| ABCD Score | **1.03** | **1.02 – 1.04** | **1.02** | **1.01 – 1.04** | **1.07** | **1.06 – 1.09** |
| Loss of function gene |  |  |  |  |  |  |
| *1 copy* | 1.02 | 0.93 – 1.11 | 1.00 | 0.92 – 1.08 | 1.02 | 0.94 – 1.11 |
| *2 copies* | 1.03 | 0.91 – 1.16 | **1.15** | **1.02 – 1.28** | **1.13** | **1.00 – 1.27** |

## Discussion

We found that the ABCD-GENE score was associated with an increased risk of vascular events among clopidogrel users in a population-based cohort in the UK. However, it was also associated with an overall increased risk of vascular events, specifically in aspirin users who do not use clopidogrel, as well as in non-users of either drug. In clopidogrel users, aspirin users, and in non-users of either drug, the clinical component, ABCD, was also associated with an increased risk of vascular events. We also found that participants with two LOF alleles in *CYP2C19* were at an increased risk of vascular events in users of aspirin and non-users of either medication but not in users of clopidogrel.

The ABCD-GENE score was developed and validated in high risk cohorts of patients undergoing percutaneous coronary intervention and was shown to be associated with both HPR and with recurrent vascular events^5^. It has also been shown to be associated with increased risk of recurrent events in a population of patients with minor stroke and transient ischemic attacks^8^. While the results from our study confirm previous findings, it also suggests that the effect of the ABCD-GENE score is not specific to clopidogrel in a large and predominantly European ancestral population.

The finding that the clinical component, ABCD, was associated with increased risk of vascular events is not surprising as the clinical components themselves – age^13^, BMI^14^, CKD^15^, and diabetes mellitus^16^ are all known risk factors for vascular events. Thus, the association of the ABCD component in clopidogrel users, aspirin users, and non-users of either drug suggests that it is the vascular risk factors that are driving the association. Several pharmacological studies have shown that these clinical factors are associated with HPR^17^, and that higher doses of clopidogrel^18^ may be needed to achieve the same platelet reactivity levels. As such, it remains uncertain how much of the increased vascular risk in patients is due to the direct impact of the vascular risk factors themselves, and how much of the increased vascular risk is due to poorer response to medications (i.e. HPR).

The number of *CYP2C19* LOF alleles was not associated with recurrent events in clopidogrel. A previous study from the UKB that focused on clopidogrel users of European ancestry did find an increased risk of vascular events amongst UKB participants with LOF alleles^19^. The inclusion of participants with non-European ethnicities in our study may have resulted in a dilution of the true effect, leading to a lack of statistical power to find an association in clopidogrel users. Additionally, we found that *CYP2C19* LOF alleles were associated with vascular events in aspirin users, and in non-users of either drug. This may be due to pleiotropy, as *CYP2C19* has been known to metabolize several classes of medications including tricyclic antidepressants, benzodiazepines, anti-infectives, and anticonvulsants^20^. The proportion of people with *CYP2C19* LOF alleles varies drastically across ancestries, with Asians having the highest proportion of LOF alleles^21^. It is uncertain whether the effect of the ABCD-GENE score would be stronger in an Asian setting rather than the predominantly European cohort in this study.

While this study benefits from a large sample size in a diverse and representative cohort comprised of individuals at variable risk for primary and secondary vascular events, we also acknowledge several limitations. Firstly, as the study population was mainly of European ancestry, the results of this study may not be generalizable to non-European populations, particularly due to differences in *CYP2C19* LOF allele prevalence across ethnicities^21^. Populations with a higher LOF allele frequency may demonstrate larger and more clopidogrel-specific effects. Secondly, as vascular events were derived from hospital admission records, we may not have captured all vascular events as some may have been treated in the outpatient sector. As such, we cannot be confident about the indication for the prescription of clopidogrel (primary prevention versus secondary prevention) and therefore cannot analyze these populations separately. Thirdly, as this was a population-based cohort, we were not able to determine platelet reactivity levels, and are therefore unable to determine whether the clinical factors (ABCD) were associated with vascular events independently or via clopidogrel drug response. Lastly, as we were using information on clopidogrel prescription, rather than consumption, we may not account for the effects of medication compliance on vascular events.

In conclusion, our study suggests that the ABCD-GENE score has utility, but is nonspecific to clopidogrel in its ability to identify patients at high risk of vascular events in a population-based cohort of primary European ancestry.

## Abbreviations

ABCD-GENE: Age, BMI, Chronic Kidney Disease, Diabetes, and Genetics
ABCD: Age, BMI, Chronic Kidney Disease, Diabetes
BMI: Body Mass Index
BNF: British National Formulary
CKD: Chronic Kidney Disease
HPR: High on treatment platelet reactivity
ICD: International Classification of Diseases
INN: International Nonproprietary Names
IS: Ischemic Stroke
LOF: Loss of Function
PCI: Percutaneous Coronary Intervention
MI: Myocardial Infarction
UKB: UK Biobank

## Acknowledgments

This research was conducted using UK Biobank Application 36993.

